# Social, economic, and environmental factors influencing the basic reproduction number of COVID-19 across countries

**DOI:** 10.1101/2021.01.24.21250416

**Authors:** Jude D. Kong, Edward W. Tekwa, Sarah A. Gignoux-Wolfsohn

## Abstract

**Objective:** To assess whether the basic reproduction number (*R*_*0*_) of COVID-19 is different across countries and what national-level demographic, social, and environmental factors characterize initial vulnerability to the virus.

**Methods:** We fit logistic growth curves to reported daily case numbers, up to the first epidemic peak. This fitting estimates *R*_*0*_. We then use a generalized additive model to discern the effects, and include 5 random effect covariates to account for potential differences in testing and reporting that can bias the estimated *R*_*0*_.

**Findings:** We found that the mean R0 is 1.70 (S.D. 0.57), with a range between 1.10 (Ghana) and 3.52 (South Korea). We identified four factors-population between 20-34 years old (youth), population residing in urban agglomerates over 1 million (city), social media use to organize offline action (social media), and GINI income inequality-as having strong relationships with *R*_*0*_. An intermediate level of youth and GINI inequality are associated with high *R*_*0*_, while high city population and high social media use are associated with high *R*_*0*_. Environmental and climate factors were not found to have strong relationships with *R*_*0*_.

**Conclusion:** Studies that aim to measure the effectiveness of interventions should account for the intrinsic differences between populations.

## Introduction

The COVID-19 pandemic, caused by the SARS-CoV-2 virus, has passed the first peak in the majority of countries in the world. Scientists, health officials and citizens have tried to anticipate and explain why the epidemic initially (i.e., before novel interventions) unfolded differently among countries, but only now has the relevant data reached sufficient global reach and temporal length to begin statistical analyses. Existing studies that examine some of the factors that may contribute to differences among countries together are generally applied to metrics such as mortality, daily and cumulative case numbers, or effective reproduction number (1–4). These metrics are time varying and sensitive to reporting and testing differences, and are therefore not easily comparable across countries. For instance, decreasing testing would allow the reported cases to drop, making raw case reporting incomparable across countries.

A key metric, *R*_*0*_, has the practical advantage of being reliably estimable (5) and comparable across countries even if testing and reporting rates are different, so long as these rates are either constant or change in roughly the same way over time. *R*_*0*_ is the basic reproduction number that indicates how many secondary infections are caused by an infected individual at the beginning of an epidemic (6). Without interventions, the portion of the population that is expected to be infected or immunized before the epidemic ends would be 1-1/*R*_*0*_. For example, an *R*_*0*_ of 3 implies that ⅔ of the population would have to be infected or immunized by the end of the epidemic. *R*_*0*_ for COVID-19 has variably been estimated between 1.4 (7) and 8.9 (8), with a likely value of 2.5 (9). Many studies either implicitly assume or are understood to imply that *R*_*0*_ is intrinsic to the infectious disease (9), but it is increasingly acknowledged that many non-interventive factors could affect heterogeneity in *R*_*0*_ among local populations or countries (10). Interventive responses that occur during the initial exponential phase of COVID-19 can be understood as proximate causes of differences in *R*_*0*_ across populations, but ultimately they are likely pre-adaptations anchored on existing social, demographic, and environmental factors. Later interventions generally affect *R*_*e*_, the effective reproduction number at any given time during the epidemic (4).

Our goal is to use a diverse and comprehensive set of demographic, social, and environmental-climatic factors to begin explaining differences in the initial dynamics of COVID-19 across countries. The predictors are non-contemporary with COVID-19, meaning they were measured before the current epidemic began. The dependent variable is the basic reproduction number *R*_*0*_, which is derived from the maximum growth rate of COVID-19 (number of additional hosts infected per infected individual per day) within a country. *R*_*0*_ can be estimated from the beginning of epidemic curves (5). The results in this study cannot be used to infer the eventual epidemic sizes among countries, which are still unfolding and can be very different from the initial dynamics due to novel interventions. We exclude proximal explanations of *R*_*0*_, such as enacted policies during the initial rise of COVID-19, because such explanations would contain statistical endogeneity - the initial epidemic growth may have partly caused the responses, therefore the responses cannot be simply used as predictors. Instead, our study focuses on how pre-existing country characteristics can explain the initial growth phases of COVID-19, although still without implying causation. We did not attempt to include all possibly relevant covariates because of high correlations even among a limited set, and because the limited number of countries dictate that a small subset should be preselected in order to retain sufficiently positive degrees of freedom for statistical analyses. Observed correlation between the covariates tested here and *R*_*0*_ may be caused by any number of other covariates that correlate with the identified covariates. Observed relationships should therefore be used for hypothesis generation and further investigation.

Covariates chosen belonged to seven categories: demographics, disease, economics, environment, habitat, health, and social. All of these categories have been suggested previously as possible factors for COVID-19 transmission. The most common factors previously studied were temperature (11–24), pollution (13,25–31), precipitation/humidity (18,32,33), population density (34,35), age structure (1,36,37), and population size (1,11,31). For these and additional covariates either previously studied or only mentioned in the media, we rely on statistics measured at a national level. A review of previously found effects on initial COVID-19 epidemic rates related to *R*_*0*_ are documented in Table 1. We examined these categories simultaneously in order to better understand which group may have a larger influence on *R*_*0*_ and should therefore be investigated further at both the national and other scales. This analysis is not meant to be exhaustive or definitive, but rather to help reveal baseline epidemiological differences across countries, shape the direction of future research on COVID-19, and understand infectious disease transmission in general.

**Table 1.** Covariates and previous findings. Data sources are cited under the covariate column. Previous effects on epidemic rates are not necessarily on basic reproduction number *R*_*0*_, but rather on cumulative case load, daily cases at certain stages, or effective reproduction number. Effects on epidemic rates are recorded as positive (+), negative (-), insignificant (0), or non-monotonic (u-shape or n-shape). Effects accompanied by (?) are theoretical.

| Category | Covariate | Previously found effects |
| --- | --- | --- |
| demographics | **Youth:** Population ages 20-35 (% of total population) (50) | (+) (36,37)<br>(-) (1) |
| demographics | **Total Pop:** Population total (50) | (+) (11,31)<br>(0) (1) |
| disease | **Mort Resp:** Mortality rate from lower respiratory infections (per 100,000) (51) | (0) (67) |
| disease | **Mort Infect:** Mortality rate from infectious and parasitic diseases (per 100,000) (51) | (-) (68) |
| economic | **GINI:** GINI index (income inequality, 100=high) (69) | (+) (61)<br>(-) (36) |
| economic | **Business:** Ease of doing business index 2019 (1=most business-friendly regulations) (70) |  |
| environmental | **Temperature:** °C January-March (71) | (-) (11–18)<br>(+) (19–22)<br>(0) (23)<br>(n-shape) (24) |
| environmental | **Precipitation:** mm January-March (71) | (+) (32,33)<br>(u-shape) (18) |
| environmental | **Pollution:** PM2.5 air pollution, mean annual exposure (micrograms per cubic meter) (50) | (+) (13,25–30)<br>(-) (31) |
| habitat | **City:** Population in urban agglomerations of more than 1 million (% of total population) (50) | (+) (34,35) |
| habitat | **Urbanization:** Urban population (% of total population) (50) | (+) (34,35) |
| health | **GHS:** Global Health Security detection index (72) | (0) (1) |
| health | **Nurses:** Nurses and midwives (per 1,000 people) (50) | (-) (1) |
| social | **Social Media:** Average People's Use Of Social Media To Organize Offline Action (4=high) (73) | (-?) (58)<br>(+?) (56) |
| social | **Internet Filtering:** Government Internet filtering in practice (4=low) (73) |  |
| social | **Air Transport:** passengers carried per capita (50) | (+) (36)<br>(0) (74) |

## Methods

All data and code are available on a Github repository (38).

### Estimating the basic reproduction number of COVID-19 among countries

The basic reproduction number *R*_*0*_ (the dependent variable) is given by the formula (39):

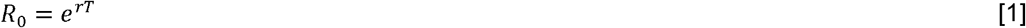

where *T* is the serial interval of COVID-19 (time delay between the symptom onset of a primary case and their secondary case) and r the initial growth rate of COVID-19. *T* has been estimated to be between 4-8 days; here we use 5.8 days (40–42). To Estimate r, we fit the rate of change in cumulative cases of a logistic growth model, with parameters *r* (intrinsic growth rate) and *K* (theoretical epidemic size without intervention), to observe time series in daily confirmed cases (5,43). The logistic growth model is superior to fitting an exponential curve to early case numbers given that case numbers do plateau in reality. In addition, the logistic growth model performs as well or better than more complicated models when confronted with data (5,44). Mechanistic models with multiple compartments (45) and with time-dependent rates (46,47), may be more realistic for COVID-19 outbreaks that in some places exhibit multiple peaks, but such models contain more parameters, require much more data, and are statistically harder to infer reliably. Such complexity is also likely not necessary to describe the initial outbreaks, which appear qualitative logistic (Figure 1).

**Figure 1.**
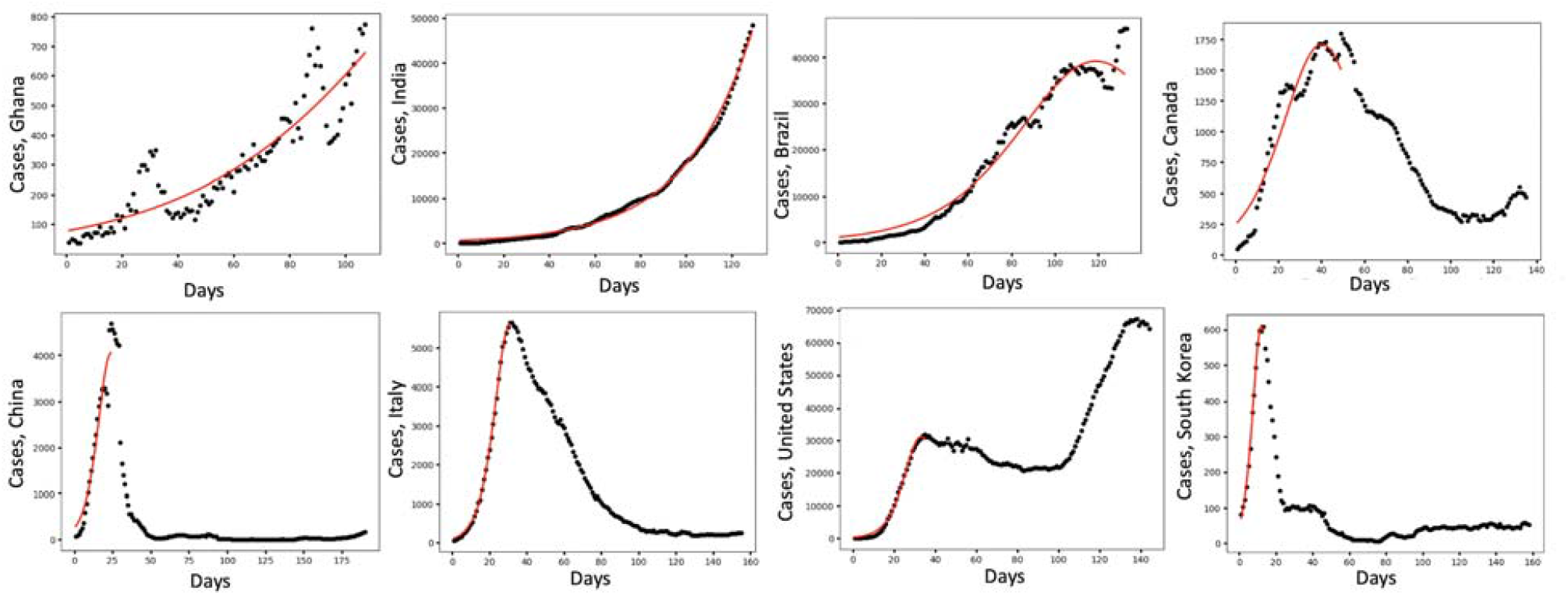
The COVID-19 daily cases. Dots represent daily cases averaged over a 7-day window, and curves are fitted based on the logistic growth model. Example countries are arranged from top left to bottom right in order of increasing basic reproduction number (*R*_*0*_).

In the logistic growth model, the cumulative case number *I* is given by:

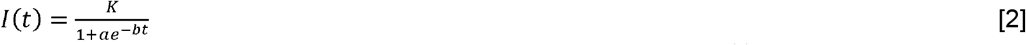

⍰ 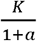 is the initial number of infected persons i.e 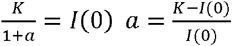
⍰ *K* is the total number of people infected at the end of the outbreak
⍰ *b* is the intrinsic or the maximum infection growth rate per infected host (growth rate for short)
⍰ 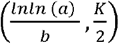 the point of maximum spread of SARS-CoV-2
⍰ *b/K* is the effective between-virus competition rate, where competition is for susceptible hosts

We truncate all COVID-19 reported daily case time series (48,49) to the day with the highest daily count, because some countries have lingered near peak daily count for much longer than a logistic growth model would predict, which would pull the model peak to later than the actual date of peak incidence and thereby underestimates *r*. We manually checked each time series and ensured that the highest daily count only occurred during a first peak. We included all countries that were at least 6 days into a period with at least 30 daily cases as of July 29, 2020, after truncating at the peak. We eliminated countries whose logistic growth model *R*^*2*^ was less than 0.9. Countries were assigned to the regions of Asia-Australia, Africa, Eurasia, Europe, Middle East, North America, and South America. Eurasia included countries that simultaneously belong to both Asia and Europe, plus Ukraine and Uzbekistan due to geopolitical proximity. The Dominican Republic was assigned to North America, while Panama was assigned to South America, as these were the only Central American countries in the final list.

Some countries do not report daily, have variable reporting delays, and may have changed reporting methods resulting in dramatic spikes in cases for particular dates. To circumvent this inaccuracy in date, we used the 7-day rolling average (right aligned) for daily cases (48,49). While this rolling average causes data from nearby dates to be autocorrelated, it should only underestimate the *p*-value of the fit but not bias the parameter estimates.

### Covariates

Next, we compiled data on predictors for each of the countries studied from seven categories (demographics, disease, economics, environmental, habitat, health, and social) from publicly available databases (Table 1). We chose covariates that are diverse, specific, and do not obviously covary; for example, gross domestic product per capita was not used because it covaries with many other more precise covariates. In addition, we chose covariates that are comparable across countries; for example, we chose nurses per capita over doctors per capita because in many countries, nurses are the primary caregivers. For each predictor, we used the most recent available data, which ranged from 2000-2019. When appropriate, data reported in absolute numbers were divided by total population to obtain per capita figures. Data with highly skewed distributions were log-transformed and all distributions were centred and standardized before regression. Four additional covariates were examined but were eliminated through sequential variance inflation factor (VIF) analysis based on the mixed effect generalized additive model described in Section 2.3 (adapted from the ‘rms::vif’ package in RStudio1.2.5033). The goal is to reduce the collinearity of the final covariate set, so that we can make better statistical attributions to how each covariate affects *R*_*0*_. In the analysis, we eliminated the covariate with the highest VIF and iterated the elimination procedure until a representative and epidemiologically reasonable set was left (the set in Table 1). The eliminated covariates were: 1. population greater than 65 years old (50), 2. life expectancy at birth (50), 3. hospital beds per capita (50), and 4. mortality rate attributed to unsafe water, unsafe sanitation and lack of hygiene (51).

### Statistical analysis

After compiling the variables, we fitted the generalized additive model (GAM) using the ‘mgcv’ package in RStudio1.2.5033, to analyze the effects of the covariates listed in Table 1, on the *R*_*0*_ value across the globe. The covariates are standardized for effect comparisons. The main advantage of GAMs over traditional regression methods are their capability to model non-linear relationships (a common feature of many datasets) between a response variable and multiple covariates using non-parametric smoothers. The general formula of a GAM is:

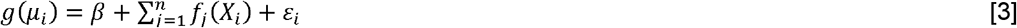

Where *g*(β_*i*_) is a monotonous link function relating the independent variable to the given covariates, β is any strictly parametric component in the model, such as intercept, *f*_*j*_(*X*_*i*_) is the variable explained by the nonparametric smoothing function, and ε_*i*_ is identically and independently distributed as a normal random variable.

Two sets of analyses are performed: 1. fixed effect model on *R*_*0*_; 2. mixed effect model on *R*_*0*_, with region, the total number of days to the first 30 cases (measured from when China had her first 30th cases), gross domestic product per capita (GDP), average under-reported percentage (52), and total number of available data points as random effects. These random effects are meant to capture differences in reporting and testing standard. GDP (50) is additionally expected to correlated with many other covariates, so using it as a random effect allows us to better understand the effects of other more precise and less aggregative metrics.

There are concerns that different COVID-19 detection capabilities among nations may affect the estimated growth rates of the disease and the regression results. Some estimates of detection differences among countries have been made (53). However, we observe that if under-reporting is constant in time within countries, then the estimated *r* and therefore *R*_*0*_ would not be affected - only *K* would be artificially depressed. On the other hand, if under-reporting is non-constant in time, then *r* would be affected (54). For example, a country that responds strongly after the arrival of COVID-19 may ramp up testing capability, which would decrease under-reporting over time. This would cause *r* fitted to the reported case data to be an overestimate. On the other hand, if a country’s detection capability erodes over time due to a shortage of test kits or a decision to stop testing non-severe cases, then *r* would be underestimated.

There are ongoing efforts to correct for these temporal biases based on delayed mortality rates (52,55), but the results are currently not credible for smaller countries with poor reporting. At this point we must rely on the reported case numbers, and use random effects to partially account for possible biases.

We use the *anova()* function in R to compare the candidate models and see which one provides the best parsimonious fit of the data. Because these models differ in the use of the random variables, ANVOA will test whether or not including random effects leads to a significant improvement over using just the given covariates without any random variables. For goodness of fits test, we use a chi-squared test.

## Results

### Basic reproduction number of COVID-19 among countries

Figure 1 and Figures S1-S4 show growth curves fit to observe time series in daily confirmed cases across countries. Figure 2 and Table S1 summarize estimated *R*_*0*_ across countries. For the countries considered, the basic reproduction number *R*_*0*_ has maximum values in South Korea, Australia and Luxemburg, with 3.52, 3.35 and 3.00 and minimum values in the Dominican Republic, Ghana, Indonesia with 1.10, 1.10, 1.11. Overall, the mean *R*_*0*_ is 1.70 with a standard deviation of 0.57. Belgium (1.71), Iceland (1.72), and Japan (1.79) are the closest to this mean *R*_*0*_.

**Figure 2.**
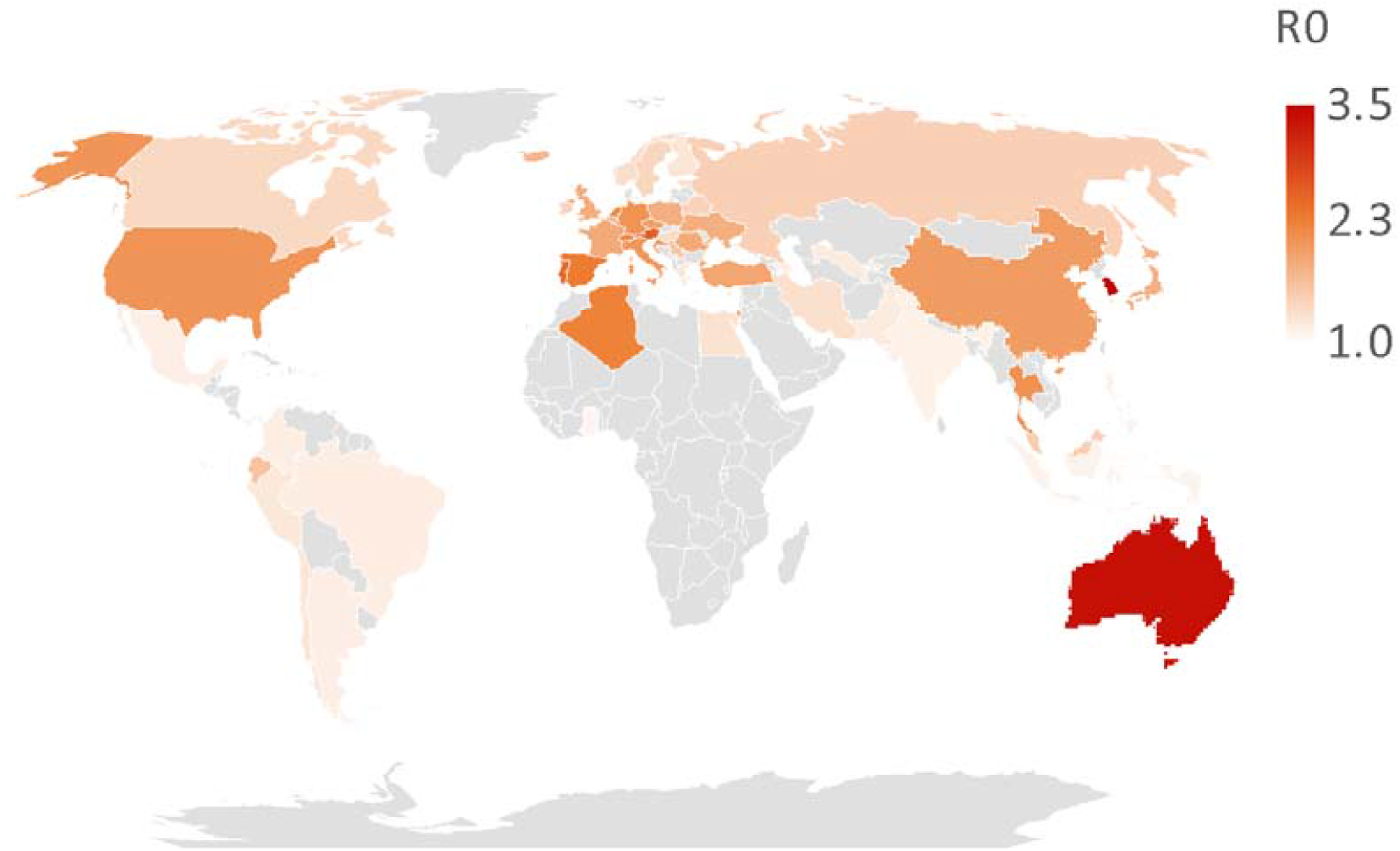
Estimated basic reproduction numbers (*R*_*0*_) for countries across the globe. Gray countries are not included in our analysis.

### Mixed effects GAM model

The explained deviance is 75.3%, this indicates that the model has a high explanatory power and predictability. The four fixed effect covariates with *p*-values below 0.1 are youth, city, social media, and GINI inequality. An intermediate value of youth (population between 20-34 years old) and GINI inequality are correlated with high *R*_*0*_. On the other hand, an intermediate level of city population (population in urban conglomerates over 1 million) is correlated with low *R*_*0*_. Finally, social media use to organize offline action is positively correlated with high *R*_*0*_ (Figure 3, Table S2). Figure 4 shows how eight exemplary countries covering a wide range of *R*_*0*_ are characterized by different demographic and social profiles. The profiles show that the countries’ ranking in covariate values mostly conform to the statistical trends suggested by GAM. For example, Ghana, with the nearly lowest *R*_*0*_, has a low portion of population in large urban agglomerates, relatively low social media usage, a large youth population, and a high GINI inequality index, which conform with the profile for low *R*_*0*_. South Korea and the United States, which have high *R*_*0*_ values, have a high portion of population in large urban agglomerates, a high social media usage, and an intermediate youth population, which conform with the profile for high *R*_*0*_. However, South Korea also has a relatively low GINI, while the United States has a relatively high GINI, whereas an average GINI is overall associated with the highest *R*_*0*_. This illustrates that countries with high *R*_*0*_ tend to fit the statistical high *R*_*0*_ profile in most but not all dimensions. Other countries examined, with lower *R*_*0*_, had profiles that diverge further from the statistical high *R*_*0*_ profile (Figure 4).

**Figure 3.**
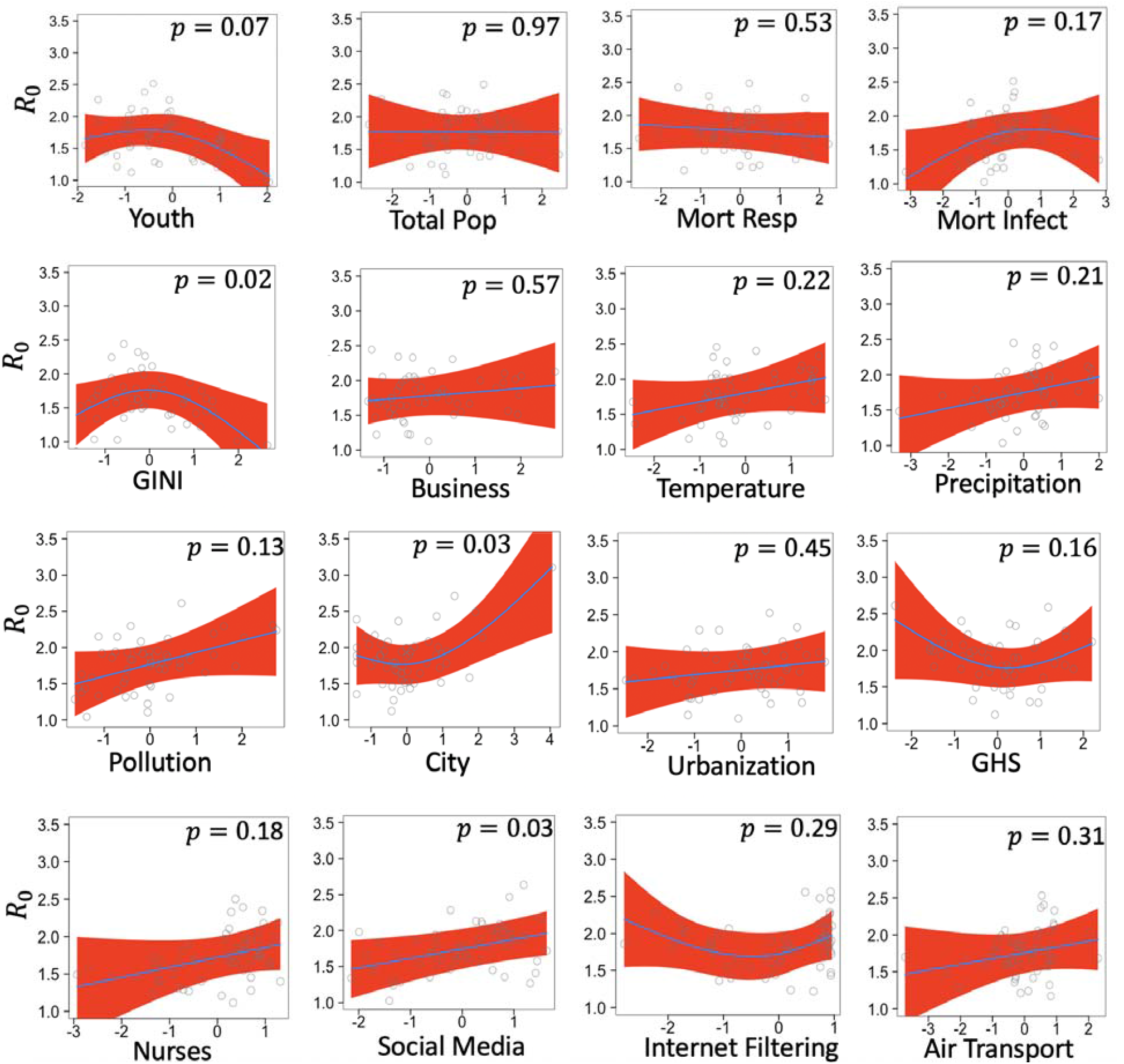
Mixed GAM derived partial effects (smoother plot) of the covariates, on *R*_*0*_. Circles are partial residuals, and red shades are 95% confidence intervals.

**Figure 4.**
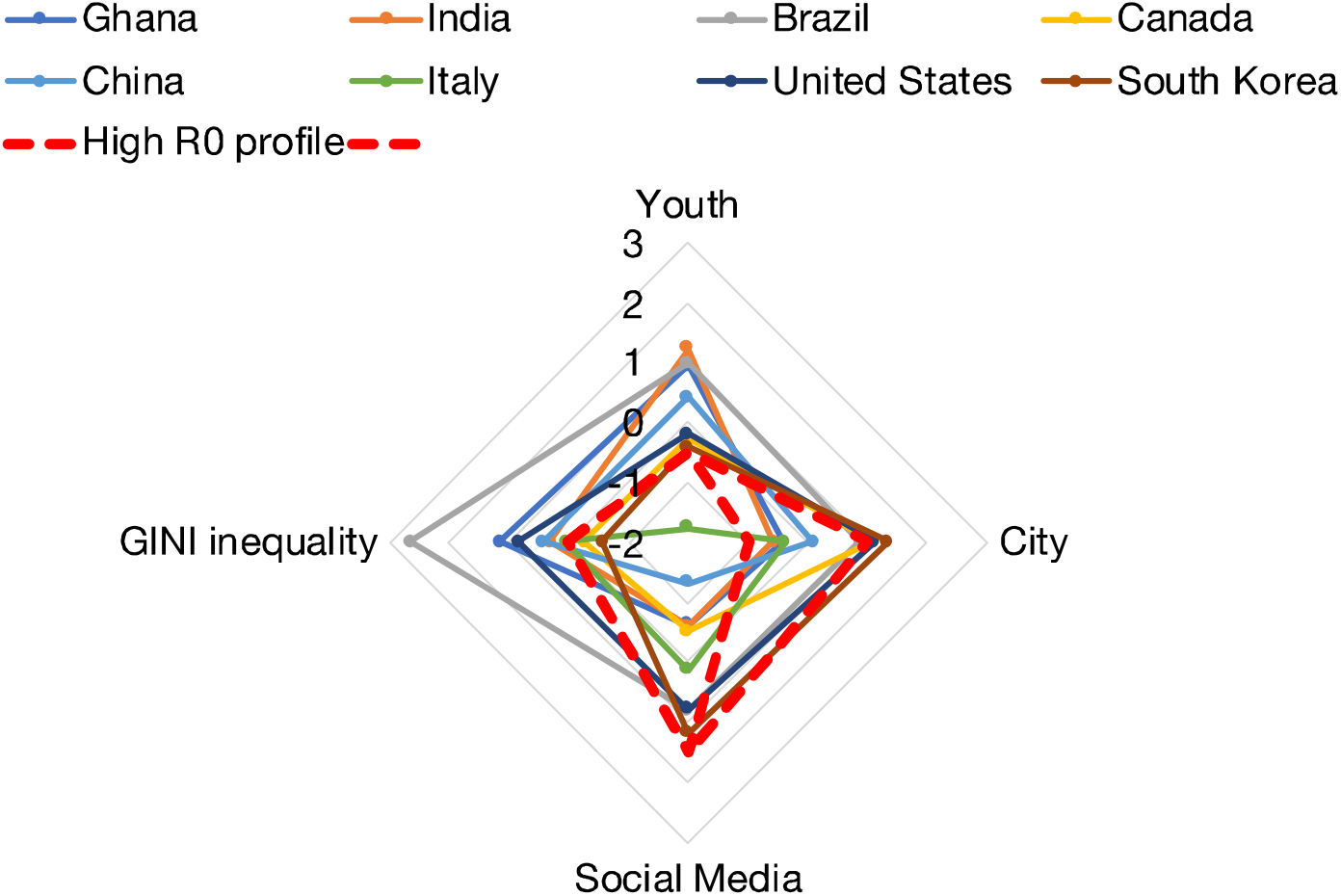
Country profiles. The four characteristics (youth, city, social media, and GINI inequality) with the lowest *p*-values in the mixed effect GAM are plotted (centred and standardized) for 8 countries representing, from top left to bottom right in the legend, increasing *R*_*0*_. Red dashed lines represent alternative high *R*_*0*_ profiles based on the mixed effects GAM model.

### Comparison with the fixed effect GAM model

For the fixed effects only model, the explained deviance is 65.5%. Figures S5 shows the effect of the covariates on *R*_*0*_, and Table S3 contains the statistical results. The ANOVA comparison of the mixed and fixed effects models shows a DF Deviance of 1.793 and *p*=0.006. This means that adding random effects to the model lead to a significantly improved fit over the fixed effects model. Some covariates have *p*-values below 0.1 in the fixed effects model but not in the mixed effects model (temperature, internet filtering). Conversely, some covariates have lower *p*-values in the mixed effects than in the fixed effects model (city, social media). These differences illustrate that random effects are important in controlling for potential biases in the raw daily COVID-19 reporting data.

## Discussion

We found that across the globe, *R*_*0*_ (1.70±0.57 S.D.) was variable and on average slightly lower than previous estimates (8,9). However, previous studies focussed on data from China and other countries with early epidemic onset, which our estimates show to have higher than average *R*_*0*_. We identified four factors (youth, city, social media, and GINI inequality) as having strong relationships with COVID-19 *R*_*0*_ across countries. Environmental factors, which are the most common factors previously identified (temperature (11–24), pollution (13,25–31), precipitation/humidity (18,32,33)), did not have strong relationships with *R*_*0*_ when other factors are considered simultaneously, although pollution, temperature, and humidity all have positive associations.

The positive relationship between social media usage and *R*_*0*_ observed here has not been previously found for COVID-19. The trend may be proximally caused by the propagation of false information on social media, for example in downplaying the potential danger of COVID-19, the effectiveness of masks and social distancing, or propping up conspiracy theories on the disease (56). One study showed that more than 80% of online claims about COVID-19 were false at the beginning of the pandemic (57). These proximal mechanisms, at least at the initial onset of COVID-19, seemed to have overridden the potential benefits of social media as an accurate information spreader that allows people to assess the true risks (58). This result may be related to the finding that in social networks false information spreads faster than the truth (59). In the initial stage of COVID-19, there was an information void regarding the nature of the disease and effective interventions, so it appears that false information filled an important void for people in countries where social media and reality were tightly weaved. However, social media may help slow a contagion’s spread once scientific information becomes available. Our result highlights the need to consider the dynamic role that social media plays in epidemics (60).

The quadratic relationships of youth and GINI inequality with *R*_*0*_ indicate a more complex underlying tradeoff than is previously appreciated, which was either monotonically positive (36,61) or negative (1,36). A large youth population may confer resilience against the disease (36,37) while also increasing the transmission rate (62); conversely an old population may be more susceptible to the disease (63,64) but exhibits a reduced transmission rate. The synergistic result is that an intermediate level of youth is related to the highest *R*_*0*_. A high GINI inequality index, referring to the amount of income inequality across a population, may mean the physical segregation of population segments and thus initially halted Covid growth across the population (36), while a low GINI may indicate better social integrations and fewer people left at high risk exposures (61). Therefore, an intermediate GINI is related to the highest *R*_*0*_. The fact that intermediate values are related to the highest, not lowest, *R*_*0*_ suggest that the hypothetical risk mechanisms - youth transmission and elder susceptibility, and social mixing and uneven risk exposures - work synergistically (rather than antagonistically) when both are present. That is, these risk mechanisms together lead to a more-than-additive increase in *R*_*0*_.

An intermediate level of city-dwelling population (population in urban conglomerates over 1 million) is related to the lowest *R*_*0*_. A high level of city dwelling is expected to increase *R*_*0*_ because of high contact rates and conforms with the main empirical trend (34). However, it is unclear why a low level of city dwelling is also associated with a high *R*_*0*_, although the rise is relatively slight. In comparison to the quadratic effects of youth and GINI inequality, the effect of city dwelling appears close to monotonic.

Our analysis is based on coarse-grained country-level case data, without explicitly correcting *R*_*0*_ estimates using temporal trends in testing, reporting, and mortality. The factors we analyzed hold across regions within a country to some extent, but it can be argued that each factor or its substitute can be measured more locally (10,65) and result in better statistical power. *R*_*0*_ can also be estimated using less phenomenological, more mechanistic models such as multiple-compartment (eg. susceptible-exposed-infectious-recovered-susceptible) (45), social network (66), or time-varying (46,47) models. However, these approaches are more data intensive and not current available in many countries. Our country-level analysis of *R*_*0*_ serves as a coarse grain baseline for future analyses pending data availability. An international perspective like the one we took here can help us understand COVID-19 in a broader context, even though we sacrifice the ability to infer local causality.

We emphasize that *R*_*0*_ is not indicative of eventual outbreak sizes or the nature of subsequent waves. Given the same population, a higher *R*_*0*_ can lead to a higher outbreak size, but this does not account for intervention measures that occur after the initial epidemic growth. For instance, a high initial epidemic growth may provide a strong signal to both citizens and governments, which then may mount a stronger response to limit the outbreak size than if the initial growth were weaker. For example, South Korea and Australia had high *R*_*0*_ (3.52, 3.35) but low cumulative case numbers (490, 1073 per million on Oct 17, 2020 (48,49)). In contrast, countries such as Brazil and Peru have low *R*_*0*_ (1.18, 1.24) and yet struggle to control the epidemic (cumulative case number=24,465, 26,156 per million on Oct 17, 2020 (48,49)). The dynamic coupling between *R*_*0*_ response is one reason why it is harder to infer the effectiveness of intervention without taking into account how pre-existing characteristics relate to initial epidemic growth. It is reasonable to believe that early interventions are actually symptoms of pre-existing social, demographic, and environmental characteristics and are not easy to implement in some countries.

The factors influencing *R*_*0*_ identified here reflect the naive or intrinsic factors that may determine a country’s vulnerability to the novel Coronavirus. While both government and citizen interventions have since been implemented in different ways, the current study can inform both the ongoing effort to control the pandemic and efforts to anticipate and control future coronavirus epidemics. The *R*_*0*_ values calculated here serve as baseline expectations for how fast COVID-19 would spread if interventions were to be prematurely lifted, given that the percent of population susceptible to COVID-19 is still relatively low. The baseline expectation for *Re* without novel interventions should be the predicted *R0* of a location given updated covariates, particularly on environmental and air transport factors that have drastically changed since the initial stage. Future studies that aim to measure the effectiveness of interventions across locations should account for intrinsic factors identified here. Otherwise, interventions in countries with intrinsically low *R*_*0*_ may be mistaken as more effective than they are, while effective interventions in countries with intrinsically high *R*_*0*_ may be regrettably ignored.

## Data Availability

All data and code are available at: https://github.com/Jdkong/COVID-19

## Acknowledgements

We are grateful for feedback from Aria Ahmad and Korryn Bodner.

## Author Contributions

J.D.K., E.W.T., and S.G.W. designed research; all authors conducted literature search and data collection; J.D.K. and E.W.T. analyzed data; and all authors wrote the paper.

## Ethics and consent

All authors have been personally and actively involved in substantial work leading to the paper, and will take public responsibility for its content.

